# Low Intensity Vibration Protects the Weight Bearing Skeleton and Suppresses Fracture Incidence in Boys with Duchenne Muscular Dystrophy

**DOI:** 10.1101/2022.04.12.22273806

**Authors:** ML Bianchi, S Vai, G Baranello, F Broggi, S Judex, TN Hangartner, CT Rubin

**Author notes:** deceased (ML Bianchi). ***ClinicalTrials.gov Identifier***: NCT05281120. ***Ethical Publication Statement:*** We confirm that we have read the Journal’s position on issues involved in ethical publication and affirm that this report is consistent with those guidelines.

## Abstract

**Introduction/Aims:** The ability of low intensity vibration (LIV) to combat skeletal decline in Duchenne Muscular Dystrophy (DMD) was evaluated in a randomized controlled trial.

**Methods:** Twenty DMD boys were enrolled, all ambulant and treated with glucocorticoids (mean age 7.6, height-adjusted Z-scores (HAZ) of hip BMD −2.3). Ten DMD boys were assigned to stand for 10min/d on an Active LIV platform (0.4g @ 30Hz), while 10 stood on a Placebo device. Baseline and 14-month BMC and BMD of spine, hip and total body were measured with DXA, and trabecular bone density (TBD) of tibia with QCT.

**Results:** All children tolerated the LIV intervention well, with daily compliance averaging 78%. At 14 months, TBD in the proximal and distal tibia remained unchanged in Placebo (−0.7% & −0.8%), while rising 4.1% and 4.5% in LIV. HAZ for hip BMD and BMC in Placebo declined 22% and 13% respectively, contrasting with no change from baseline (0.9% and 1.4%) in LIV. Fat mass in the leg increased 33% in Placebo, contrasting with 20% in LIV subjects. Across the 14-month study, there were four incident fractures in three placebo patients (30%), with no new fractures identified in LIV subjects.

**Conclusions:** These data suggest that non-invasive LIV can help protect the skeleton of DMD children against the disease progression, the consequences of diminished load bearing, and the complications of chronic steroid use.

## Introduction

Duchenne muscular dystrophy (DMD) is an X-linked recessive genetic disease due to a mutation in the dystrophin gene. DMD is characterized by severely reduced or absent dystrophin in skeletal muscles, with progressive muscle degeneration and fibro-fatty replacement.[1] Affected individuals experience progressive muscle weakness beginning in early childhood, with loss of ambulation by the age of 12 years [2] and reduced life expectancy due to the progressive cardio-respiratory impairment.[3] While there is no cure, the introduction of glucocorticosteroids (GCs) in the 90’s have been shown to partially preserve muscle strength, protect pulmonary function, delay onset of ambulatory decline, reduce severity of scoliosis, and extend life.[4] GC treatment of DMD is now initiated at a much earlier age, as built on clear evidence that the sooner they initiate the treatment the greater its effectiveness in slowing progression of the disease,[5, 6] and with ventilator support life expectancy can approach 40y.[7]

In concert with GC treatments, novel dystrophin restoration therapies have slowed muscle collapse, and have mitigated - to a limited degree - some functional decline of DMD patients,[8] yet concomitant skeletal complications persist.[9] Indeed, compromised bone mineral density (BMD) and bone mineral content (BMC), are considered primary causal factors in increased occurrence of fragility fractures.[10, 11] As compared to healthy age-matched boys, DMD patients show a significant disparity in bone quantity and quality in the lower limbs, which in part is a consequence of reduced weight-bearing and muscular activity on bone [12] associated with this crucial period of growth.[13]

During the period that DMD boys retain their ability to walk, lumbar spine BMD is only slightly decreased but then drops precipitously when they lose ambulation.[14] In some contrast, lower limbs are more severely affected, reflected by reduced BMD and BMC at hip even while walking ability is only slightly impaired. DMD boys have 30% less trabecular bone density in the tibia than healthy controls, falling below 50% on loss of ambulation.[15] This decline in bone quality is accompanied by an increase in fracture risk: Individuals with muscular dystrophies are at a 1.4-fold increased risk of fracture when compared with population-based controls, a risk that rises with age and triples when glucocorticoids have been used for at least six months.[16] These fractures have a devastating impact: 4 out of 9 ambulatory DMD boys never recover to walking status after a fracture.[14] Any strategy that protects bone will help reduce fractures and prevent early loss of ambulation.

It is well known that physical activity is critical in achieving and maintaining bone mass accrual across the years of growth in children.[13] To a degree bone mass can be correlated to muscular strength during adolescence,[17, 18] while animal studies show that lean muscle mass persists as a significant determinant of bone quantity and quality even when challenged by a dystrophic phenotype.[19] Although the decline in bone status in DMD parallels deterioration of muscle phenotype, it does not appear to be directly caused by the disease itself but instead as a secondary consequence of the reduced loading that parallels muscle weakness.[1, 20]

Exercise regimens as a strategy to preserve muscle function and bone accrual in DMD children shows some promise, however, it is still unclear if this can protect from skeletal decline or if instead can lead to an increased risk of fractures.[21] As a surrogate for exercise, low magnitude mechanical signals, delivered using Low Intensity Vibration (LIV), have been demonstrated to have an anabolic effect on bone in animal models,[22] driven by some degree by biasing bone marrow mesenchymal stem cells away from adipogenesis and towards osteoblastogenesis.[23] High-frequency, low magnitude mechanical signals are omnipresent in the functional load regime,[24] arising from the dynamics of muscle contraction.[25] It is proposed here that introducing these signals as a surrogate for exercise could help protect the skeleton of DMD patients, who, because of muscle decline, have reduced muscle-induced loading of bone. The present study is aimed at evaluating the safety, tolerability and effects on bone status of LIV in children with DMD.

## Methods

### Study Design

The study was originally designed as a prospective, double-blind, randomized, placebo-controlled 12-month trial on 20 boys with DMD. Due to a severe illness of the principal clinician (MLB) at the 1-year mark, protocol duration was extended to 14-months. The study was approved by the Ethical Committees of both institutions involved in subject recruitment (Istituto Auxologico Italiano IRCCS and Istituto Neurologico Besta IRCCS) and conducted according to the Clinical Good Practice rules. Recruitment for the trial was initiated before the FDAAA 801 requirement for registration with clinicaltrials.gov, so retroactive registration was made after the trial was completed (identifier: NCT05281120). Considering that all potential subjects were minors, informed consent and assent was obtained from the boys’ parents.

Each enrolled subject received a LIV platform to take home. Participants were randomized to either receive an Active or Placebo platform. Subjects, parents, and study team were blinded to the Active/Placebo status of the device. Subjects and their parents were instructed to use the platform for 10 minutes each day, scheduled at any time during the day convenient for its use. Instructions included to stand upright, in a relaxed stance, wearing only socks to cover the feet. The patients were provided a diary to record the day, time, and minutes of their platform use, and they were instructed to record any day where the LIV treatment was not used, a strategy that had shown close similarity to electronic recording of compliance in prior clinical trials.[26, 27] Patients in both the LIV and Placebo groups received weekly phone calls from the study’s clinicians and hospital staff to reinforce their commitment to participating.

### Subjects

Recruitment for the study was initiated through DMD family groups with clinical histories at the two enrolling institutions. Twenty ambulant boys diagnosed with DMD, aged 4-15 years (mean age 7.6±3.9 years), were enrolled. Baseline data are reported in Table 1. Inclusion criteria included: diagnosis of DMD; ability to stand up and walk (some balance assistance allowed, but full weight-bearing necessary); treatment with a fixed dose of prednisone (1.25 mg/kg every 2 days, according to the treatment protocol of the Istituto Neurologico Besta IRCCS); treatment with 25-D (calcifediol, 0.7 mcg/kg/d); and dietary calcium intake equal to the internationally recommended daily allowance (RDA). All inclusion criteria had to be met for at least 6 months before starting the study. DMD diagnosis was made at the Muscle Pathology and Immunology Unit of the Istituto Neurologico Besta IRCCS. The diagnostic criteria were based on clinical data, molecular analysis, morphological evaluation, and/or immunochemical analysis confirming the absence of dystrophin in muscle fibers.

**Table 1:** Baseline Characteristics of Body Habitus and Bone Density.

|  | Placebo | Active | P-value |
| --- | --- | --- | --- |
| Age (yrs.) | 6.6 ± 1.6 | 9.4 ± 3.1 | <0.05 |
| Weight (kg) | 20.8 ± 4.6 | 31.5 ± 10.2 | 0.06 |
| Height (cm) | 114.0 ± 12.5 | 127.4 ± 15.7 | <0.05 |
| Height (Z-score) | 1.1 ± 1.0 | 1.3 ± 1.4 | 0.45 |
| Hip BMD (g/cm<sup>2</sup>) | 0.530 ± 0.071 | 0.503 ± 0.073 | 0.52 |
| Hip BMD HAZ | -1.39 ± 1.25 | -2.59 ± 0.62 | 0.06 |
| Hip BMC (g) | 6.90 ± 2.09 | 8.69 ± 4.26 | 0.41 |
| Hip BMC HAZ | -2.08 ± 1.28 | -3.12 ± 1.12 | 0.21 |
| TBLH BMD (g/cm<sup>2</sup>) | 0.529 ± 0.053 | 0.596 ± 0.080 | 0.08 |
| TBLH BMD HAZ | -0.33 ± 1.05 | -1.24 ± 0.96 | 0.10 |
| TBLH BMC (g) | 422.5 ± 98.7 | 544.5 ± 198.7 | 0.16 |
| TBLH BMC HAZ | -1.07 ± 0.63 | -2.03 ± 0.67 | <0.05 |
| Proximal Tibia Trabecular Bone Density (mg/cm<sup>3</sup>) | 240.2 ± 61.2 | 184.9 ± 46.3 | 0.10 |
| Distal Tibia Trabecular Bone Density (mg/cm<sup>3</sup>) | 203.8 ± 33.1 | 173.5 ± 16.1 | <0.05 |
| Tibia Cortical Bone Density (mg/cm<sup>3</sup>) | 1227.1 ± 40.4 | 1245.3 ± 58.1 | 0.44 |

### LIV Platform

The 20 enrolled DMD boys were randomized into two groups and assigned to either an active LIV platform delivering 0.4g (where 1g is Earth’s gravitational field or 9.8m/s^2^), with a 30Hz (cycles per second) sinusoidal vibration (N=10) or Placebo platform (N=10), which – to mask status of the device - emitted a 500Hz hum through an onboard loudspeaker but produced no vibration signal through the plantar surface of the standing child. Peak to peak accelerations of 0.4g at 30Hz require displacements of less than 120 microns, or the thickness of two human hairs. 100% compliance would be the use of the platforms for 10 minutes each day, seven days per week, across the length of the study. To avoid over-use, the device was restricted to a maximum of 10 minutes of LIV within any calendar day. A LIV prototype designed for adults weighing between 40 and 115kg was modified for use by children by reducing spring constants of the springs that suspend the top platen, allowing a weight range of 15-65kg.[28]

The design of the LIV platform uses closed-loop acceleration feedback to drive an electromagnetic actuator, ensuring a high-fidelity sinusoidal signal,[29] a design which can safely deliver these barely perceptible mechanical signals to standing subjects.[30] Signals at this frequency and intensity are considered a non-significant risk by the FDA,[31] and as defined by the International Standards Organization Advisory 2631, exposure to vibration signals at this frequency and magnitude is considered safe for up to four hours of exposure each day.[32]

### Evaluation Program

At baseline and end-of-protocol, all enrolled DMD boys underwent the following evaluations: weight, height, fracture history and an interview with a dietician to evaluate the daily intake of calcium. Weight was measured with an electric scale to the nearest 0.1 kg. All patients were able to stand up (with aid in some cases), and the standing height was measured with a stadiometer.

Baseline and end-of-protocol DXA and QCT bone imaging studies were performed at Istituto Auxologico Italiano IRCCS (Milan, Italy). At baseline only, bone age was evaluated with hand radiography. All the enrolled subjects underwent neurological functional evaluation at baseline and end-of-protocol at the Istituto Neurologico Besta IRCCS, as part of their standard clinical monitoring.

### Dietary Calcium Intake

Calcium intake was evaluated at baseline, administered by the same skilled dietician. Each subject had a 20-30-minute dietary interview, including a food frequency questionnaire and 24-h dietary recall, administered in a private room in the presence of one or both parents. In addition, the nutritional composition of the lunch menus (5 days/week) from the children’s schools were evaluated. The frequency consumption (daily, weekly, and monthly) of each food item was evaluated. For each item, the children indicated the size of their usual meals using photographs of small, medium, and large portions. The food frequency questionnaire included 16 main food groups (e.g., milk and dairy products; pasta and rice; drinks; cereals and oven products such as bread and biscuits, etc.), classified according to nutrient composition and customary use by Italian children.[33] The calcium content of water (tap and bottled mineral water) was considered, obtaining the calcium content of local tap water at the patients’ locations, or from the labels of the mineral waters. The analysis was performed using an Italian National Institute of Nutrition software (Winfood 1.0b), providing detailed food composition data.[34]

### Vitamin D and Bone Formation Markers

From all subjects, at both baseline and 14 months, blood samples were collected between 8-9a.m., after an overnight fasting. Biochemical measurements of serum 25-hydroxy vitamin D (25-D), 1,25-dihydroxy vitamin D (1,25-D), parathyroid hormone (PTH), bone-specific alkaline phosphatase (BSAP) and osteocalcin (OC) were performed. 25-D was quantified by radioimmunological assay (RIA; DiaSorin); intra- and inter-assay coefficient of variation (CV) 3.5% and 7.5%; 1,25-D by radio receptor assay (Nichols Institute Diagnostics, San Juan Capistrano, CA, USA) intra- and inter-assay CV 5.6% and 7.9%, PTH by immunoradiometric assay (IRMA; DiaSorin Inc, Stillwater, MN, USA) intra- and inter-assay CV 2.8% and 4.7%; and OC by RIA; (Technogenetics, Milano, Italy); intra- and inter-assay CV 3.6% and 6.9%.

### Computed Tomography

Quantitative computed tomography (QCT) scans of the tibia were performed with a GE QCT (34 slices) at both legs with the same protocol in all boys. One of the coauthors (TH) analyzed these data, blinded, with special software.[35]

### Bone Mineral Density

Bone mineral content (BMC) and density (BMD), as well as fat and lean mass, were measured by dual energy x-ray absorptiometry (DXA; Hologic Discovery Horizon A densitometer) at lumbar spine, proximal femur, and total body. At Istituto Auxologico Italiano IRCCS, a strict DXA quality control procedure, including the instrument’s daily phantom calibration, is standard and was regularly followed during the study. The DXA CV, with repositioning, was 0.62-1% for spine and 0.64-1.09% for total body, depending on age. Total body BMC, BMD, fat and lean mass were calculated excluding head (TBLH), the most appropriate measurement in a growing skeleton,[36] considering the different patterns of cranial development.[37] Height-Adjusted Z-scores (HAZ) for BMD and BMC for spine, hip and TBLH were calculated based on healthy boys of the same age.[38]

Two *post-hoc* analyses were performed. First, changes in BMD and BMC in the spine and hip were normalized to BMD/BMC of TBLH, and second, normalized to BMD/BMC of the arm, as automatically segmented from the total body DXA measures. As the LIV platform challenges only the weight-bearing bones and is not considered a ‘systemic’ stimulus to the skeleton,[31] BMD and BMC of TBLH and the segmented changes in the arm were considered an intra-subject measure that could be used to approximate DMD-driven changes in the status of the skeleton that occurred across the 14 months of study, with less ‘exposure’ to the LIV signal. As normalization of hip or spine to TBLH would include the very regions that were being examined (i.e., the hip being included in TBLH would mask changes in the hip), normalization to the arm would exclude those regions being evaluated (i.e., the hip would be assessed relative to a change in the arm, independent of the hip).

### Fractures

Fracture history, including circumstances, skeletal site, date, and type of intervention, was taken at baseline and updated at each clinical visit. All fractures were documented by radiography. At baseline and end-of-protocol, lateral radiographs of thoracic and lumbar spine were taken to evaluate the presence of vertebral fractures.

### Statistical Analyses

Data were expressed as the mean ± SD. For group comparisons, Student’s *t* test for unpaired (Placebo vs LIV) and paired (Baseline vs 14-months) samples were used, as appropriate. For intragroup comparisons, changes at 14-months were expressed as absolute and relative differences from baseline values. Chi-square tests were used to determine whether occurrence of fractures was significantly different between Placebo and LIV groups. Statistical significance was defined at p<0.05. Because of the small sample size, p-values between 0.05 and 0.20 were reported as potential differences (trends).[39] P-values of greater than 0.2 represented a lack of statistically significant differences.

## Results

Due to investigator illness at the one-year mark of the first subject, the intended 12-month protocol was extended to 14 months (∼400 days), a delay incorporated into the follow-up schedules of all boys. All enrolled patients completed the 14-month study, and all subjects tolerated the treatment well and declared they had been happy to use the LIV platforms. Neither subjects nor parents reported any discomfort, inconvenience, or adverse effects.

### Baseline Characteristics

Sixty-six patients were screened for the study, with 37 not meeting inclusion criteria and nine who declined to participate (**Figure 1**). Across the twenty enrolled subjects (**Table 1**), diagnosis of DMD was made at 2.8 + 1.2 y, with steroid therapy begun at 5.1 + 1.1 y. There were significant differences in age between Placebo (6.6 + 1.8 y) and LIV (9.4 + 3.4 y) subjects (p<0.05). There were also significant differences in height between Placebo (114.0 + 12.5 cm) and LIV (127.4 + 15.7 cm) subjects (p<0.05), and a trend in difference in weight between Placebo (20.8 + 4.6 kg) and LIV (31.5 + 10.2 kg) subjects (p<0.20). Height Z-score for Placebo (−1.11 + 1.0) was not different than LIV (−1.3 + 1.4). While assignment of platform status was blinded, these disparities in age, weight and height remain a major limitation of the study and are addressed in the discussion.

**Fig. 1.**
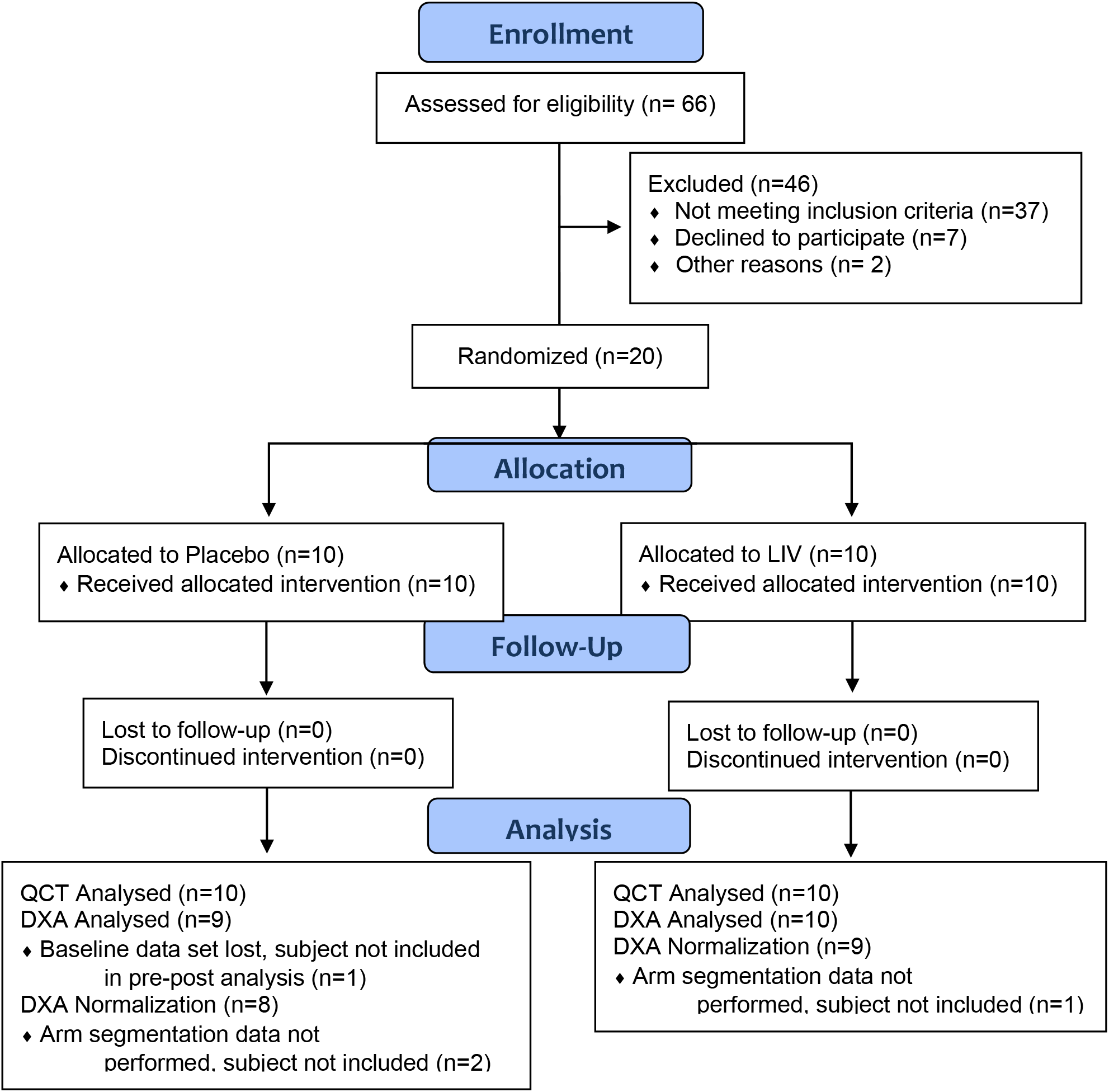
Consort Diagram for the Duchenne Low Intensity Vibration trial.

The compliance was evaluated using the patients’ diaries and the platforms’ records was 79% in LIV and 75% in Placebo groups. Specific compliance measures per subject, including logbooks, were lost to follow-up, and thus efficacy as a function of compliance could not be determined.

### 14-Month Changes in Body Habitus

When considering subject specific changes in height from baseline, there was a 5.6% increase in the Placebo group (p<0.05), as compared to a 4.5% increase in height in the LIV group (p<0.05), but no difference in growth rates between the groups (p=0.27). When considering subject specific changes in weight from baseline, there was a 13.1% increase in Placebo (p<0.05) as compared to a 17.8% increase in the LIV group (p<0.05), representing a 5.8% difference in body mass gained between the groups (p<0.20).

### Calcium and Vitamin D Intake

The dietary calcium intake was 650 ± 132 mg/day at baseline and 710 ± 129 mg/day at month 3, as established by phone interview, measures in line with the average calcium intake of healthy Italian children.[34] After dietary adjustment at month 3, it increased to 1,186 ± 233 mg/day by end of study (p<0.05 vs. baseline). The adherence to diet and vitamin D intake was estimated via interviews with children and parents about calcium intake from foods, by measuring the children’s’ serum 25-D levels, and by checking the used calcifediol bottles that the patients were asked to keep and bring back. “High adherence,” defined as taking at least 80% of the prescribed doses, was estimated in 73 ± 6.3% of patients, and even those with lower adherence had an increased calcium intake with respect to baseline (average increase: 280 ± 110 mg/day). High adherence to calcifediol treatment was estimated in 84 ± 5.3% of patients. There was no difference between LIV and Placebo groups in any dietary measures (data not shown).

### 14-Month Changes in BSAP, PTH and OC and Vitamin D

Baseline and 14-month markers of bone turnover are summarized in **Table 2**. Follow-up serum measures were lost to follow-up for five subjects in each group, so that baseline/end-of-study comparisons are based on n=5 in each group. Baseline BSAP in the Placebo group did not change at 14 months (+1.8%; p=0.27), while LIV rose 7.2% (p<0.20). Baseline PTH in the Placebo group did not change at 14 months (p=0.47), while rising 12% in LIV (p<0.035). Baseline OC did not change at 14-months in either the Placebo (−2.1%; p=0.40) or LIV group (+16.9%; p=0.33). Baseline 1,25-D did not change at 14-months in either the Placebo (−10.5%; p=0.24) or LIV group (+ 6.6%; p=0.32). Baseline 25-D increased 31% in Placebo (p<0.20), but did not change in LIV (−5%; p=0.42)

**Table 2:** Bone Turnover Markers.

|  | Placebo |  | Active |  |
| --- | --- | --- | --- | --- |
|  | Baseline | 14 months | Baseline | 14 months |
| BSAP | 51.6 ± 17.9 | 50.7 ± 23.3 | 61.2 ± 35.2 | 65.6 ± 22.3 |
|  | -1.8%; p=0.27 |  | +7.2%; p<0.20 |  |
| PTH | 23.8 ± 7.5 | 24.1 ± 1.0 | 29.2 ± 8.2 | 32.7 ± 10.4 |
|  | <1%; p=0.45 |  | +12%; p<0.05 |  |
| OC | 61.7 ± 9.6 | 60.4 ± 18.9 | 77.3 ± 32.0 | 90.4 ± 37.9 |
|  | -2.1%; p=0.40 |  | +16.9%; p=0.33 |  |
| 1,25-D | 37.7 ± 11.6 | 33.8 ± 11.5 | 51.5 ± 11.0 | 54.9 ± 12.4 |
|  | -10.5%; p=0.24 |  | +6.6%; p=0.32 |  |
| 25-D | 17.5 ± 7.8 | 23.0 ± 9.8 | 38.0 ± 17.6 | 36.0 ± 10.0 |
|  | +31.4; p<0.20 |  | -5.3; p=0.42 |  |

### QCT Measures of Bone Density in the Tibia

Data are reported as n=10 in each group. After 14 months, trabecular bone density (TBD) in the proximal region of the tibia remained unchanged in the Placebo group (−0.7%; p=0.44), while the 4.1% rise in the LIV group trended towards significance (p<0.20). TBD in the distal region of the tibia remained unchanged in the Placebo group (−0.8%; p=0.43) while rising 4.5% in the LIV subjects (p<0.20, **Figure 2**). When considering subject-specific changes from baseline, TBD of the distal region was 4.9% higher in LIV as compared to the Placebo group (p<0.20). Increases in cortical BMD of the tibial midshaft were significant in Placebo subjects (+1.8%, p<0.20) but failed to reach significance in LIV (+1.7%, p=0.55). There was no difference in changes in cortical bone in the Placebo as compared to LIV group (p=0.47).

**Fig. 2.**
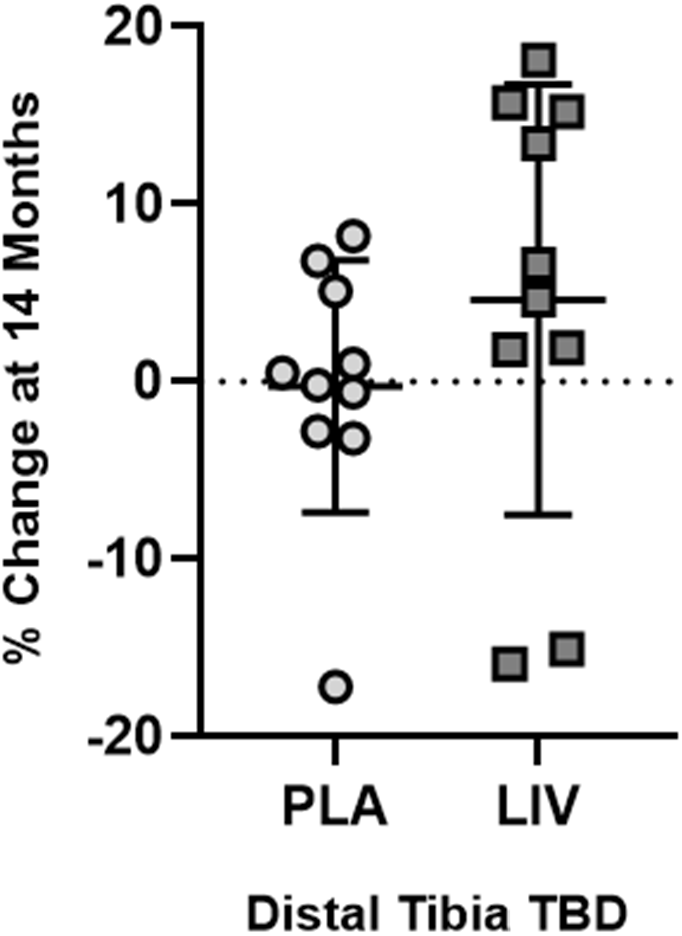
QCT measured changes (mean + SD) at 14-months in trabecular bone density in distal tibia. TBD did not change in Placebo (circles, 0.8% below baseline, p=0.43), in some contrast with TBD increases typical to healthy boys of the same age.[76] LIV increased TBD by 4.8% (squares, p<0.20 from baseline).

### BMD and BMC Measures of TBLH

Data reported are n=9 for both LIV and Placebo; baseline data sets could not be located for one subject in each group. As measured by DXA, BMD changes in TBLH in the Placebo group showed a 3.5% increase over the course of the 14-month period (p<0.05), as compared to a 2.8% increase in the LIV group (p<0.05). Changes in the two groups were not different from each other (p=0.44). BMC changes in TBLH in the Placebo group showed a 15.6% increase over the course of 14 months (p<0.05), as compared to a 13.1% increase in the LIV group (p<0.05). Changes in the two groups were not different from each other (p=0.39).

The degree to which the skeleton of these DMD subjects is compromised becomes evident with direct comparisons to age- and height-matched healthy boys. HAZ of TBLH BMD in the Placebo group were −0.3 at baseline and fell to −0.8 by the end of the 14-month protocol (p<0.05). LIV subjects at baseline were −1.2 and fell to −1.7 by the end of the experimental period (p<0.05). When evaluating absolute change in HAZ, Placebo fell by 0.52 (p<0.05), similar to the 0.51 absolute decrease measured in LIV subjects (p<0.05), with the changes between groups not different from each other (p=0.95, **Figure 3**). Absolute decreases in BMC were −0.09 in both Placebo and LIV groups, with no differences from baseline or between groups.

**Fig. 3.**
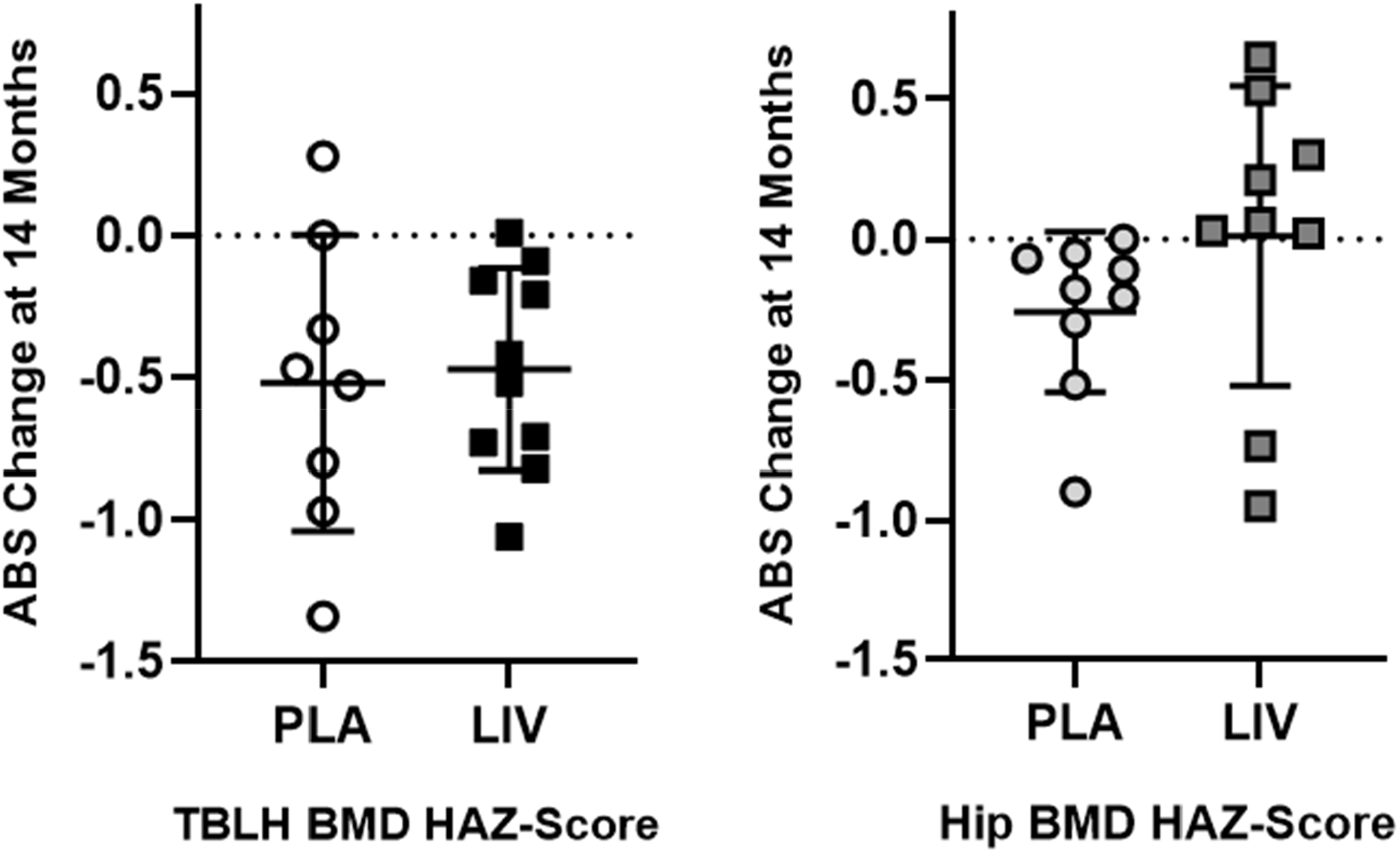
Absolute change in TBLH (left) and Hip (right) BMD HAZ score from baseline (mean + SD). The 14-month decrease from baseline in TBLH was significant for both Placebo and LIV (p<0.5), but the differences between groups was not different (p=0.95). In contrast, the absolute decrease in HAZ hip BMD was significant in Placebo (p<0.05), but there was no change from baseline in LIV.

### BMD and BMC Measures of Hip

DXA measures of BMD in the hip of Placebo subjects did not change over the experimental period (+0.6%; p=0.35), while increasing 3.0% in the LIV group (p<0.20). There was no difference between groups (p=0.31). DXA measures of BMC in the hip increased 11.4% in Placebo (p<0.05) and increased 12.6% in LIV subjects (p<0.05). There was no difference between groups (p=0.31).

### BMD and BMC Height Adjusted Z-Scores of Hip

Matched to healthy boys, hip BMD HAZ-scores of Placebo subjects at baseline were −1.4, falling 21% to −1.7 by the end of the 14-month protocol (p<0.05). HAZ-scores of Hip BMD in LIV subjects at baseline were −2.6, with a 0.9% drop at 14-months not significantly different from baseline (p=0.91, **Figure 3**). HAZ of hip BMC in the Placebo group fell 13% (p<0.20) from −2.0 to −2.3, while the baseline measure of −3.2 in the LIV group was not different at 14 months (p=0.87). Absolute HAZ in hip BMD fell 0.3 in Placebo (p<0.05) but remained similar to baseline in LIV (−0.02; p=0.45, **Figure 3**). Absolute HAZ in hip BMC fell 0.28 in Placebo (p<0.20) but remained similar to baseline in LIV (−0.04; p=0.87).

### BMD and BMC Measures of Spine

DXA measures of BMD in the spine of Placebo subjects increased 7.0% (p<0.05), while increasing 7.8% for LIV subjects (p<0.05). Changes in the two groups were not significantly different from each other (+1.5%; p=0.34). DXA measures of BMC in the spine of Placebo subjects increased 8.8% (p=<0.20), whereas BMC in the LIV group increased 13.1% (p<0.05). Changes in LIV were not different than the Placebo group (+4.3%; p=0.29).

### BMD and BMC Height Adjusted Z-Scores of Spine

Matched to healthy boys of the same age, baseline HAZ of BMD spine in the Placebo group was −0.11 at baseline, and did not change by the end of the 14-month protocol (+0.38; p=0.29). At baseline, HAZ-scores of spine BMD in LIV subjects was −0.8, and had not changed by the end of the 14-month protocol (−0.2, p=0.54). HAZ of spine BMC in the Placebo group at baseline was −1.5, and had not changed by the end of the 14-month protocol (−14%, p=0.29). HAZ of spine BMC in the LIV subjects began at −2.1, with no change at 14 months (p=0.89).

### Bone, Lean and Fat Measures of Arm

BMD changes in the left arm of the Placebo group showed a 5.9% increase over 14 months (p<0.20), as compared to a 2.9% increase in the LIV group (p=0.20). Changes in BMD in the left arm measured in the Placebo group was not different than the LIV group (p=0.49). BMC changes in the left arm in the Placebo group showed a 14.2% increase over the 14 months (p<0.05), as compared to a 10.4% increase in the LIV group (p<0.05). Over the 14-month protocol, BMC in the left arm of the Placebo group increased 6.9% more than the LIV group (p=0.20).

Fat mass in the arms of the Placebo group increased 18.2% across the 14 months (p<0.05), as compared to 15.1% in the LIV group (p<0.05). Group specific changes were not different from each other (p=0.57). Lean mass in the Placebo group increased 10.3% across the 14-month period (p<0.05), in contrast to 6.4% increase in the LIV group (p<0.05). Group specific changes were not different from each other (p=0.55).

### Fat and Lean Mass of TBLH and Leg

DXA-measured increase in TBLH fat mass across the 14 months were not different (p=0.94) between Placebo (+26.9%, p<0.05 from baseline) and LIV (+27.1%, p<0.05 from baseline). Fat mass of the left leg showed a 32.9% (p<0.05) increase in Placebo subjects as compared to a 19.8% (p<0.05) increase in LIV subjects, a 40% suppression of fat accumulation (p<0.20; **Figure 4**).

**Fig. 4.**
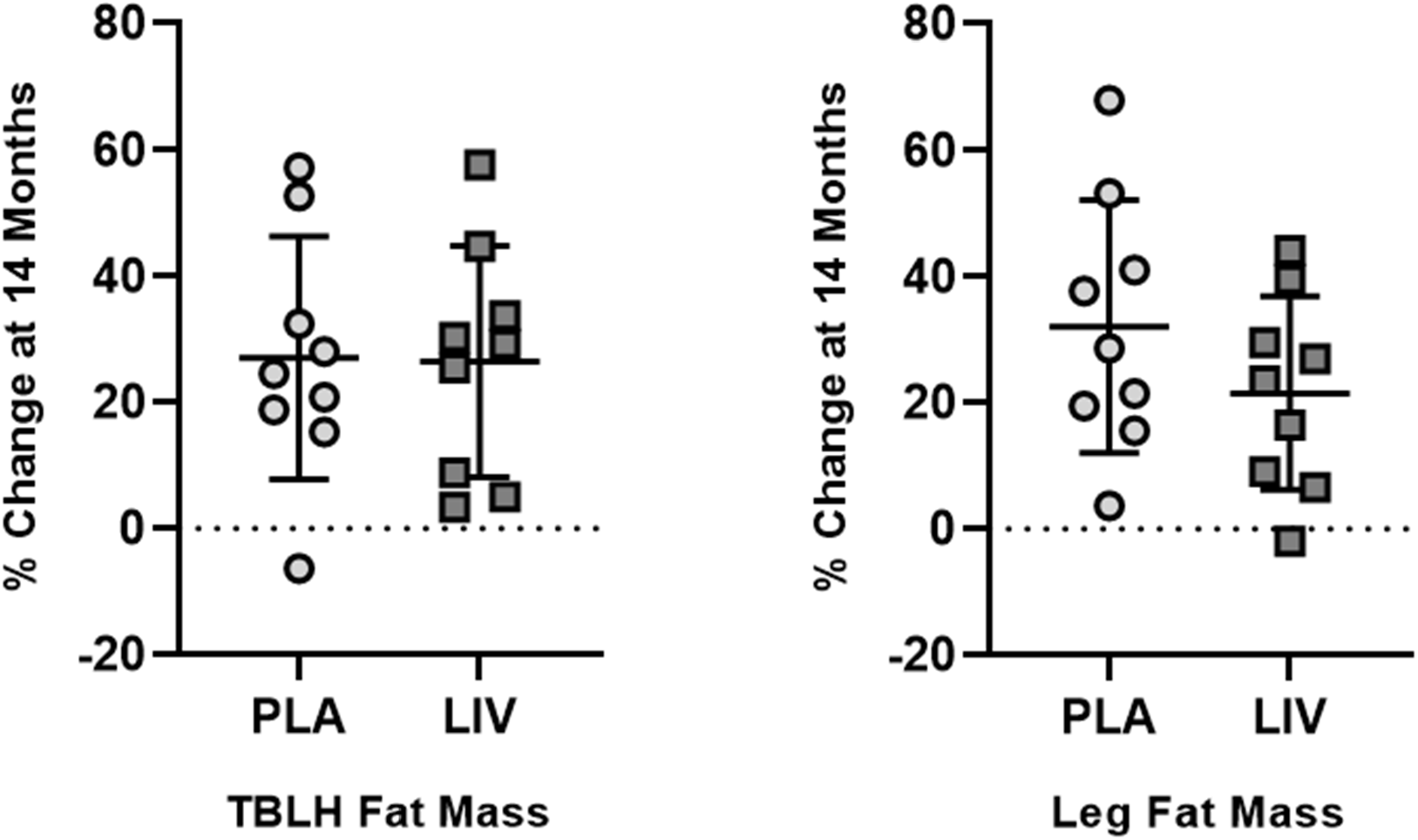
Percent increase (mean + SD) from baseline in fat mass for TBLH (left) and Hip (right). The 14-month increase from baseline in TBLH was significant (p<0.05) for both Placebo (circles) and LIV (squares), but the differences between groups was not different (p=0.95). DXA measured fat mass of left leg showed increases in both Placebo (circles; 32.9%) and LIV (squares; 21.6%) DMD subjects. Subject-specific changes showed a 40% suppression of fat mass in LIV as compared to Placebo (p<0.20).

DXA-measured increase in TBLH lean mass across the 14 months was similar (p=0.69) between Placebo (+10.7%, p<0.05 from baseline) and Active (+8.9%, p<0.05 from baseline). Lean mass of the left leg closely tracked TBLH, with a 9.6% (p<0.05) increase in Placebo subjects as compared to an 8.0% (p<0.05) increase in the Active subjects (p=0.75 between groups).

### Spine and Hip BMD and BMC Normalized to TBLH

*Post-hoc* analyses of DXA data were first performed by normalizing subject-specific parameters to that individual’s changes measured across the entire body minus the head (TBLH). This normalization was performed to determine if ‘systemic’ changes in bone quantity and quality caused by the disease, as represented by the entire skeleton (including hip and spine), would help determine if LIV influenced those regions of the skeleton that were subject to the mechanical signal delivered primarily to the weightbearing bones.[28] When normalized to changes in TBLH, BMD in the spine of Placebo subjects was not different to baseline (+2.4%; p=0.42), as compared to a 4.3% increase in LIV subjects (p<0.20). There were no differences between groups (p=0.34). When normalized to changes in TBLH, BMC in the spine dropped 6.2% (p<0.05) in the Placebo group, as contrasted to a 1.0% decrease in the LIV subjects (p=0.39). When considering subject-specific changes of BMC of spine from baseline, there was a 5.6% positive swing from the Placebo group to the LIV group (p<0.20).

Relative to TBLH, BMD in the hip fell 3.1% (p<0.05) in the Placebo group, while remaining unchanged from baseline in the LIV subjects (−0.7%; p=0.40). Group specific changes were not different from each other (p=0.43). Normalizing BMC changes in TBLH to those measured in the hip showed a 6.5% drop in BMC in the Placebo group (p<0.20) as compared to no change from baseline in LIV subjects (−1.6%; p=0.78). Group-specific changes were not different from each other (p=0.24).

### Spine and Hip BMD and BMC Normalized to Arm

*Post-hoc* analyses of DXA data were also performed where subject specific parameters were normalized to that subject’s changes measured in the arm. This normalization was performed to determine if ‘systemic’ changes in bone quantity and quality caused by the disease, as represented by an upper extremity (i.e., minus hip and spine), would help determine if LIV influenced those regions of the skeleton that were subject to the mechanical signal delivered to the weightbearing bones.[28] When normalized to changes in the arm, BMD in the spine of the Placebo subjects was not different from baseline (+1.5; p=0.52), as compared to a 4.9% (p<0.20) increase in the LIV subjects (**Figure 5**). Group-specific changes were not different from each other (p=0.21). When normalized to changes in the arm, BMC in the spine dropped 5.6% (p<0.20) in the Placebo subjects, as contrasted to a 4.8% increase in the Active group (p<0.20), a 10.4% positive swing from Placebo to Active (p<0.05; **Figure 6**).

**Fig. 5.**
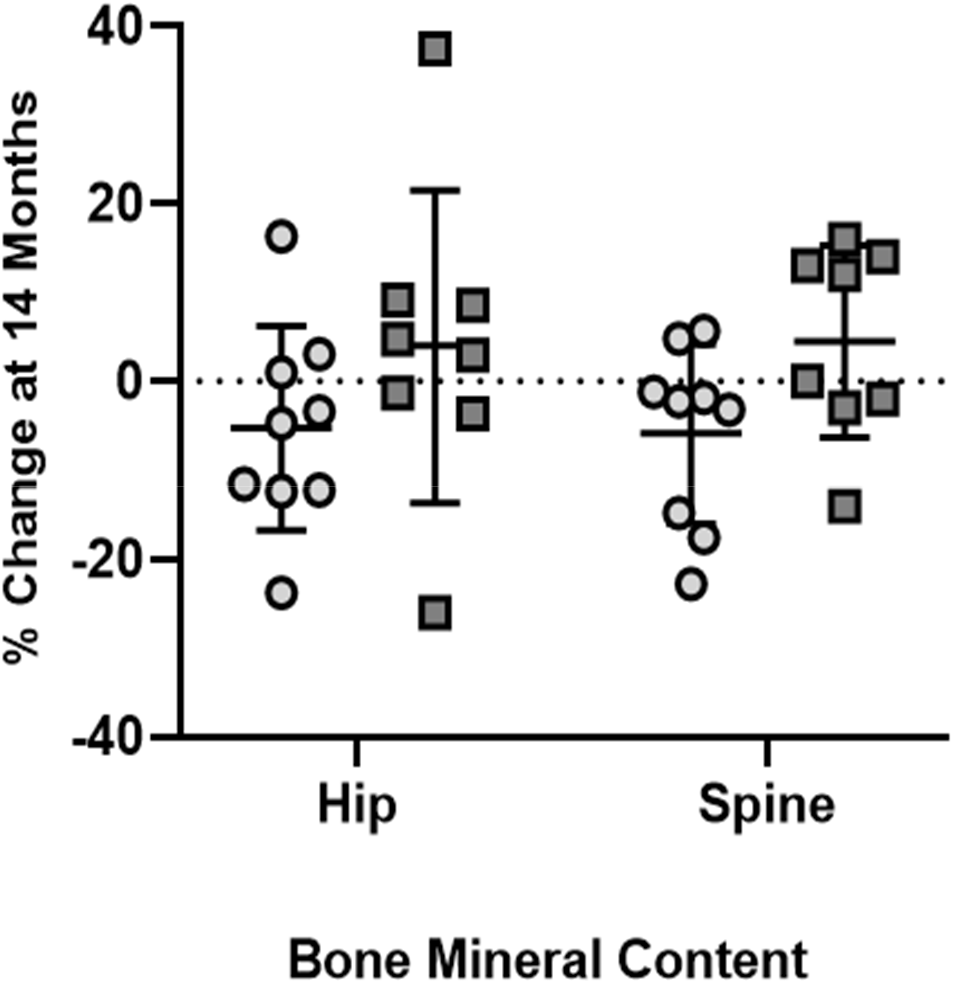
DXA measured percent change (mean + SD) in BMC at Hip and Spine at 14-months w/placebo (circles) and LIV (squares) subjects, as normalized to changes in arm (intra-subject control). Hip BMC in placebo decreased by 5.2%, while LIV increased by 4.0%, a 9.2% shift (p<0.20). BMC in spine dropped 5.9% in placebo, and increased 4.4% in LIV, a 10.2% shift (p<0.05).

**Fig. 6.**
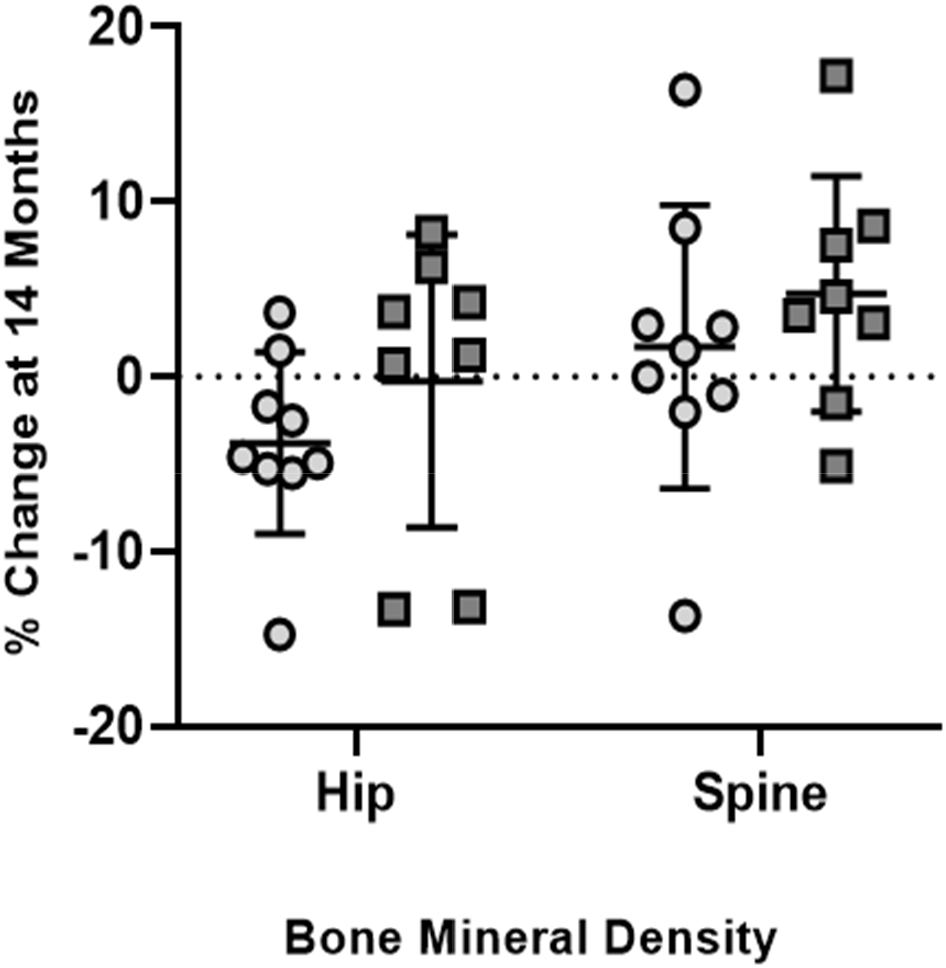
DXA measured percent change (mean + SD) at 14-months in BMD in Placebo (circles) and LIV subjects (squares), as normalized to changes in arm (intra-subject control). Hip BMD in the placebo decreased by 3.8%, while LIV remained unchanged (0.02%), a 3.5% shift (p<0.20). BMD spine increased 1.7% in placebo, and 4.7% in LIV, a 3.0% shift (p<0.20).

Normalizing BMD changes relative to those realized in the arm across 14-months to those measured in the hip showed a 4.0% fall in the Placebo subjects (p<0.05), while remaining unchanged from baseline in LIV subjects (+0.2%; p=0.80), representing a 3.5% difference groups (p<0.20; **Figure 5**). BMC in the hip normalized to that measured in the arm, there was a 6.0% drop in the Placebo subjects (p<0.20), in contrast to no change from baseline in LIV subjects (+3.7%, p=0.26), a 9.7% positive swing from Placebo to Active groups (p<0.20; **Figure 6**).

### Fractures

At baseline, across the whole group, 13 lower-limb fractures had been sustained by 8 DMD subjects before starting the study (40%). No vertebral fractures were reported in the clinical records, but baseline lateral spine radiographs revealed 6 prior fractures of dorsal vertebrae in 4 DMD subjects (1 fracture each in 3 patients; 3 fractures in 1 patient).

Regarding incident fractures at study end (month 14), there was 1 dorsal vertebral fracture and 3 appendicular fractures (2 foot fractures, 1 fibula fracture) identified in 3 patients in the Placebo group (30%), while no new fractures were identified in the LIV group, a significant difference between groups (p<0.05; **Figure 7**). The cause of these fractures is not known.

**Figure 7.**
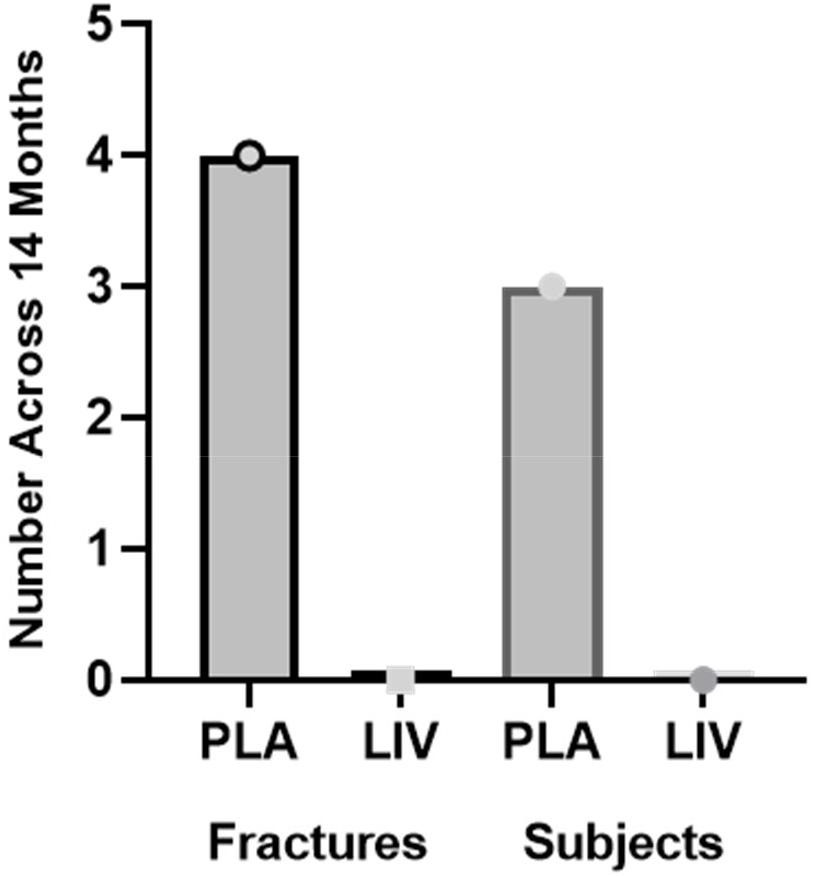
Absolute number of incident fractures (left) and subjects with fracture (right) identified across the 14-month protocol in both Placebo and LIV group (N=10 subjects in each group). While the causes of the fractures are not known, the difference between groups is significant (p<0.05).

## Discussion

Implementation of exercise programs to protect the musculoskeletal systems of children with Duchenne Muscular Dystrophy (DMD) has been complicated by fears that high-intensity or eccentric exercise may accelerate muscle deterioration, that active weight-bearing protocols are often beyond reach of this population, or – paradoxically - that these activities may increase risk of fractures.[40] As a surrogate for exercise protocols, this prospective randomized double-blind placebo-controlled study reports on the effects of LIV on skeletal bone status in children with DMD. The 20 DMD boys enrolled in this study were all ambulant, treated with the same GC regimen, with dietary calcium intake according to the RDA for age, and receiving calcifediol (25-OH vitamin D_3_) supplementation. This study showed that LIV was safe and well tolerated, as daily compliance was high and there were no reported adverse events. The study, while small, provides some insight into the potential of exogenously delivered, low-magnitude mechanical signals as a means of protecting the skeleton in an at-risk population.

LIV has been shown to increase bone mass and quality in children with disabling conditions, including cerebral palsy[41] and adolescent girls with idiopathic scoliosis.[42] LIV augments bone accretion in survivors of childhood cancer[43] and patients with Crohn’s disease.[44] In a one-year study on young women (age 15-21) with osteoporosis, LIV was shown to be anabolic to both femur and spine, as well as paraspinous musculature, representing improvements relative to that measured in placebo control and achieved while suppressing fat formation in the torso.[45] Translating LIV to children with muscular dystrophies, a one-year pilot trial evaluated LIV as a protective influence on muscle function in five patients with DMD or Becker muscular dystrophy, with the first six months exposing each subject to 10min/d of LIV (0.4g @ 30Hz), with the second six months halting the LIV intervention.[46] Timed motor function and lower extremity muscle strength remained unchanged or slightly improved during the intervention phase, but was followed by marked deterioration once LIV was discontinued. While the investigators concluded LIV to have a “stabilizing effect on lower extremity muscle function,” no measures on the skeletal system were performed.

In the DMD study reported here, changes from baseline to 14-month DXA measures of bone mineral content and bone mineral density in TBLH, arm, hip and spine showed significant increases in both the Active and Placebo groups, emphasizing that the skeleton of these children continued to grow. Despite differences in ages of the two groups, 14-month increases in BMD and BMC measures of TBLH and arm were highly similar between LIV and Placebo subjects, suggesting that skeletal growth in the non-weightbearing regions progressed in a similar fashion. Nevertheless, even with increases in BMD and BMC in LIV and Placebo subjects, comparative metrics of DMD bone quality and quantity – as established by HAZ - fall well short of that observed in healthy children, indicating that the DMD skeleton is more susceptible to fracture,[47] a risk that becomes more severe with age and extended GC use.[48]

HAZ provide information about the skeleton relative to an age, height and gender-matched average established in healthy boys. For example, the Hip BMD HAZ for this DMD cohort shows the baseline measures for LIV subjects being at −2.45, more than two standard deviations below what might be expected at that age and for that height. And while HAZ declined 21% in the Placebo group, pointing towards an escalating susceptibility to fracture, the Active group remained unchanged from baseline, pointing to a potential for LIV to limit bone loss in the weightbearing skeleton of extremely high-risk DMD patients. These regional Z-scores reinforce prior findings that skeletal quality in the lower appendicular skeleton is lower than that of the spine or total body,[49] but are encouraging in that LIV suppresses further decline in these regions of greatest risk.

When considering the influence of LIV on the skeleton as measured by DXA, trends showed increases at both the hip and spine, in both BMD and BMC, in the LIV vs the Placebo group, but none of these measures reached significance. However, when DXA outcomes were normalized, first to the overall index of TBLH, and then to 14-month changes measured in the arm, the consequences of DMD to the weightbearing skeleton became more apparent within the Placebo group, with neither the hip nor spine ‘keeping up’ with BMC and BMD increases in upper regions of the body. This failure of the weight-bearing bones to parallel increases in the arm is – at least in some part – a consequence of diminished functional demands made to these regions: as the disease progresses, those individuals with DMD are less active and not loading their skeleton in the same way that healthy boys might do.[21] Thus, while genetic programming drives the skeleton to continue to grow across adolescence,[36] robustness, the added anabolic benefit of mechanical loading, is less evident, resulting in a mismatch between bone quantity and quality in the upper and lower extremities.[14]

Normalizing bone quality and quantity in the hip and spine of LIV subjects to either TBLH or the arm suggest that these mechanical signals could serve as a protective agent, particularly when compared to the regional disparities identified in the Placebo group. For example, when normalized to specific changes in TBLH occurring over the 14 months, BMC in the spine increased by 1% in the LIV group, but fell 6.2% in the Placebo subjects, representing a 5.6% ‘benefit’ of LIV even when considering this older cohort is at greater risk. Of course, TBLH *includes* the spine and hip, and thus any changes in these regions of interest would be masked by this ‘aggregate’ measure. Normalizing solely to the arm showed that the spine in Placebo subjects had decreased 5.6% 14-months later, while there was a 4.8% increase in LIV subjects, reflecting a 10.2% benefit of low-magnitude mechanical signals. These data reinforce several important elements of mechanical loading. First, the hip and spine of Placebo subjects fall more quickly than the upper regions of the body, emphasizing that, as the disease progresses, the lower extremities and axial skeleton are more susceptible to decline due to a reduction in normal functional loading.[14] In some contrast, when BMC and BMD changes in hip and spine in the LIV group are normalized to TBLH or the arm, there is a relative increase in those measures in the weightbearing regions, suggesting that the low magnitude mechanical signals are – to a degree - successful in restoring an anabolic response in the bone tissue, adapting to a surrogate of functional load bearing. It is important to emphasize, however, that the results reported here, while encouraging, must be viewed with caution, as several are evident only after normalizing to other regions of the skeleton. It is also important to note that such a site-specific assessment is limited to interventions such as LIV, as it is a regional signal to the weight-bearing skeleton, not a systemic stimulus to the entire skeleton.

In considering LIV’s ability to protect the skeleton, it is also important to note that this approach has failed in some trials. Two years of LIV stimulated no response in the skeleton of persons of advanced age, with subjects averaging 82y.[50] However, it is also known that bone tissue in ‘older’ skeletons is not as sensitive to mechanical stimuli as in a young population.[51] Whether that is a consequence of a reduced bone cell population in aged bone,[52] that the cell population is less responsive,[53] or that bone marrow shifts towards a fat phenotype,[54] and thus less progenitors are available to recruit, is unclear, but these and other factors attenuate the adaptive response to loading and conspire to an increased risk of fracture.[55]

Prior studies of using high intensity whole body vibration (WBV; >5.0g) in DMD have not shown responses in bone, perhaps because these studies were short in duration (4, 8 & 12 weeks).[56–58] In an 8w study on 14 DMD patients and 8 with spinal muscular atrophy (SMA), high-intensity WBV for 3 minutes per day, 5x/w, aimed to improve muscle strength and function. Mild functional improvements were observed, including a significant improvement in the 6-min walking test in children with SMA.[57] No skeletal measures were made. A 12w study examined the influence of high-intensity WBV delivered 2x/w on muscle and bone in 6 ambulatory DMD patients,[56] but no significant changes in bone mass or strength were measured. Finally, 4w of high-intensity WBV, delivered 3x/w, in 4 DMD patients showed the subjects tolerated the intervention, but no significant changes in functional mobility were identified.[58] While the magnitude of the vibration in these studies are up to 20x higher than those used here (e.g., 8.0g vs. 0.4g), it is not clear that a benefit of higher intensity ultimately outweighs the added risk.[46] The safety of chronic exposure to high intensity vibration must also be considered,[59] particularly in those with skeletons at high risk of fracture.[60] Thus, while ‘magnitude’ *per se* does not seem to be a critical variable, delivering these signals each day, and for an extended period, appears to be important to harness the response of the skeleton.

As measured by DXA, there were significant 14-month increases in fat mass in TBLH and the arms of both LIV and Placebo subjects, but no difference of this increase between groups, suggesting LIV had no influence on fat phenotype in these non-weightbearing regions of the body. In some contrast, DXA pointed to a 33% increase of fat mass in the leg of Placebo subjects, 60% greater than that measured increase in LIV subjects. Thus, while fat mass increases were significant in the leg of both LIV and Placebo subjects, it was also evident that the Placebo group rose at a higher rate, a predictor of regional functional decline.[61] And while 14-month increases in lean mass of the leg were also significant in both LIV and Placebo subjects, the 2% difference between groups was not different. Importantly, fat encroachment into muscle is a major complication in DMD, starting in the lower limbs as early as the age of five,[62] compromising both function and regenerative capacity.[63] While this study was not structured to examine either the fat or muscle phenotypes *per se*, there is certainly evidence that inactivity is permissive to increased fat production in growing children,[64] that GC can promote adipogenesis and suppress osteoblastogenesis in DMD patients,[65, 66] and that LIV – like exercise[55] - has suppressed systemic adipogenesis, including subcutaneous, visceral, and marrow fat production.[23, 67]

As a more precise assessment of bone quality, QCT measures of trabecular bone density of the proximal and distal tibia showed that the Placebo group remained essentially unchanged over the course of 14-months. These data suggest that, despite increases in overall BMD and BMC as measured by DXA at the hip, spine and TBLH, reflecting growth patterns in these children, the structural elements of the bone are not keeping pace, similar to findings reported in other disabling conditions in children.[68] Whereas Placebo subjects show no change from baseline, LIV subjects show an increase in TBD across the 14 month period, suggesting that the anabolic potential of LIV is reinforcing trabecular structures in this region, similar to that measured in children with Crohn’s disease.[44] Recent findings, using MRI as an assay of bone quality, show that LIV enhances bone mineral density in the weight-bearing skeleton of post-menopausal women as well as trabecular connectivity and bone microstructure.[69] Perhaps such improvements translate to a more robust skeleton and an overall decrease in susceptibility to fracture.[70]

There is a large fracture burden in children with DMD. Results reported from the NorthStar data base show that – over a 4y period – incident fractures occurred in 28% of the 564 participants, but this rate almost doubled in those patients taking corticosteroids.[48] While GC use has shown great benefit to skeletal muscle mass and function in DMD,[71] consequences of long-term use include accelerated decline of the skeleton and greater risk of fracture. Indeed, 30% of the subjects in the Placebo group suffered an incident fracture over the 14-month period of the study. In some contrast, despite HAZ-scores well below those of the Placebo group, there were no incident fractures reported in the LIV cohort. It is entirely possible that no fractures in LIV subjects is a coincidence, or perhaps the influence of these mechanical signals serves to improve bone microarchitecture, and thus provides a proportional benefit to fracture resistance.[69] Further, as Petryk demonstrated an improvement of muscle function and strength through LIV, perhaps these small contributions work synergistically, through stability, balance and skeletal strength, to resist fractures.[46] Regardless, any means of reducing fracture incidence can have a tremendous impact on retaining quality-of-life, as over 40% of ambulatory DMD patients who suffer a fracture never return to weight bearing activities.[14]

This double-blind, prospective trial was designed to determine if LIV could effectively suppress the deleterious changes in the skeleton seen in DMD. Several limitations must be considered in interpreting the results. First and foremost, the study was designed such that the Active/Placebo devices were randomly assigned. Un-blinding revealed that the mean age of the LIV group was 3y older than that of the Placebo group. Whereas such disparities are always a risk in a small trial,[39] the observation that older DMD subjects in the LIV group were building bone and suppressing fat production can also be considered reassuring, as these boys – *by any perspective* – would be less active, at greater fracture risk, and at a later stage of disease progression. Further, reporting the 14-month outcomes in relation to HAZ-scores, helps diminish age-specific differences between groups, particularly as the BMD/BMD decreases in TBLH were identical between LIV and Placebo. Using each subject as their own control, normalizing first to TBLH and then to changes in the arm, allows for determining the relative changes to the weight-bearing skeleton as contrasted with the bones in the upper extremity, which is less ravaged by the disease.[12] Most importantly, as the fracture incidence in LIV was significantly lower than Placebo, despite a higher age and lower baseline BMD, is encouraging.

LIV is only a potential surrogate for exercise, not a replacement.[55] LIV’s efficacy is stronger in some populations than others,[72] with those that are ‘least’ responsive tending towards older cohorts, including the frail elderly.[50] That the DMD group is young, however, can only potentiate LIV’s ability to combat skeletal decline.[41–45] It is also important to emphasize that the focus of this study was bone, and not muscle, the primary system ravaged by DMD. Further studies are necessary to determine if – and how – LIV can protect muscle strength and function in the human. Prior work in the mouse has shown that LIV can stimulate hypertrophy in muscle,[73] and can promote power output in muscle units,[74] while satellite cells, compromised by endocrine deficiency, are protected by LIV.[75] Nevertheless, until clinical studies are performed, it is impossible to project whether LIV can guard the musculoskeletal ‘system’ against LIV.

Functional load bearing is essential to preserving a robust skeletal system. A devastating consequence of diseases such as Duchenne Muscular Dystrophy is that, as the anabolic element of muscle loading declines across childhood, the skeleton’s risk of fracture increases, placing quality of life at risk; over 40% of ambulatory DMD boys never recover to walking status after a fracture. Results from this 14-month study suggest that daily LIV treatment is safe, well tolerated, and may benefit the quantity and quality of the weight bearing skeleton of DMD children, and thus reduced their risk of fracture.

### In Memoriam

It is with a great sense of sorrow that we must write that the lead clinical investigator, and first author of this study, Professor Maria Luisa Bianchi, passed away in September 2020. She was an internationally renowned expert on pediatric metabolic bone disease, fully committed to clinical research that could ultimately help her patients and families of those children. We are grateful for her leadership and enthusiasm for fostering this study forward, and hope that, in some small way, this work contributes to her lasting memory and enormous impact on the science and community of pediatric endocrinology.

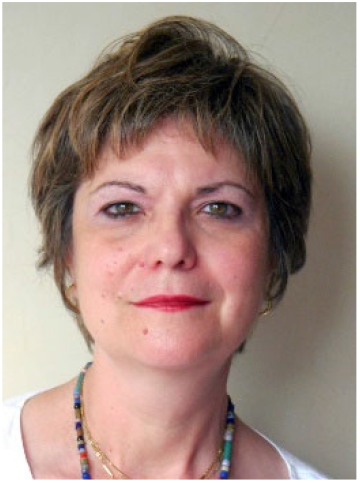

## Data Availability

All data produced in the present study are available upon reasonable request to the authors

## Acknowledgments

The authors thank the participants and their parents for their courage and commitment to participating in this study. We are hopeful that the time invested by the children and families will help the biomedical community better understand strategies to help those afflicted with this disease. We also thank Claudio Cappuccino, who was instrumental in collecting and compiling the data following the passing of his wife, Professor Bianchi. Finally, we are grateful to the Istituto Auxologico Italiano IRCCS, Milan, Italy, for providing funding for the study, and to Mr. Ian Cutts, COPA Healthcare, for the generous loan of the prototype LIV devices.

## References

1. Summer SS, Wong BL, Rutter MM, Horn PS, Tian C, Rybalsky I, et al. Age-related changes in appendicular lean mass in males with Duchenne muscular dystrophy: A retrospective review. Muscle Nerve. 2021;63(2):231–8. Epub 2020/10/27. doi: 10.1002/mus.27107. PubMed PMID: 33104257.

2. Deconinck N, Dan B. Pathophysiology of duchenne muscular dystrophy: current hypotheses. Pediatr Neurol. 2007;36(1):1–7. Epub 2006/12/13. doi: 10.1016/j.pediatrneurol.2006.09.016. PubMed PMID: 17162189.

3. Sussman M. Duchenne muscular dystrophy. J Am Acad Orthop Surg. 2002;10(2):138–51. Epub 2002/04/04. doi: 10.5435/00124635-200203000-00009. PubMed PMID: 11929208.

4. Szabo SM, Salhany RM, Deighton A, Harwood M, Mah J, Gooch KL. The clinical course of Duchenne muscular dystrophy in the corticosteroid treatment era: a systematic literature review. Orphanet J Rare Dis. 2021;16(1):237. Epub 2021/05/24. doi: 10.1186/s13023-021-01862-w. PubMed PMID: 34022943; PubMed Central PMCID: PMCPMC8141220.

5. Beytia Mde L, Vry J, Kirschner J. Drug treatment of Duchenne muscular dystrophy: available evidence and perspectives. Acta Myol. 2012;31(1):4–8. Epub 2012/06/05. PubMed PMID: 22655510; PubMed Central PMCID: PMCPMC3440798.

6. Matthews E, Brassington R, Kuntzer T, Jichi F, Manzur AY. Corticosteroids for the treatment of Duchenne muscular dystrophy. Cochrane Database Syst Rev. 2016;(5):CD003725. Epub 2016/05/06. doi: 10.1002/14651858.CD003725.pub4. PubMed PMID: 27149418.

7. Landfeldt E, Thompson R, Sejersen T, McMillan HJ, Kirschner J, Lochmuller H. Life expectancy at birth in Duchenne muscular dystrophy: a systematic review and meta-analysis. Eur J Epidemiol. 2020;35(7):643–53. Epub 2020/02/29. doi: 10.1007/s10654-020-00613-8. PubMed PMID: 32107739; PubMed Central PMCID: PMCPMC7387367.

8. Takeda S, Clemens PR, Hoffman EP. Exon-Skipping in Duchenne Muscular Dystrophy. Journal of neuromuscular diseases. 2021. Epub 2021/06/29. doi: 10.3233/JND-210682. PubMed PMID: 34180420.

9. Annexstad EJ, Bollerslev J, Westvik J, Myhre AG, Godang K, Holm I, et al. The role of delayed bone age in the evaluation of stature and bone health in glucocorticoid treated patients with Duchenne muscular dystrophy. Int J Pediatr Endocrinol. 2019;2019:4. Epub 2020/01/01. doi: 10.1186/s13633-019-0070-0. PubMed PMID: 31889957; PubMed Central PMCID: PMCPMC6927168.

10. Larson CM, Henderson RC. Bone mineral density and fractures in boys with Duchenne muscular dystrophy. Journal of pediatric orthopedics. 2000;20(1):71–4. Epub 2000/01/21. PubMed PMID: 10641693.

11. Bianchi ML, Morandi L, Andreucci E, Vai S, Frasunkiewicz J, Cottafava R. Low bone density and bone metabolism alterations in Duchenne muscular dystrophy: response to calcium and vitamin D treatment. Osteoporosis international. 2011;22(2):529–39. Epub 2010/05/12. doi: 10.1007/s00198-010-1275-5. PubMed PMID: 20458570.

12. Bianchi ML, Mazzanti A, Galbiati E, Saraifoger S, Dubini A, Cornelio F, et al. Bone mineral density and bone metabolism in Duchenne muscular dystrophy. OsteoporosInt. 2003;14(9):761–7.

13. Gabel L, Macdonald HM, Nettlefold L, McKay HA. Physical Activity, Sedentary Time, and Bone Strength From Childhood to Early Adulthood: A Mixed Longitudinal HR-pQCT study. Journal of bone and mineral research :. 2017;32(7):1525–36. doi: 10.1002/jbmr.3115. PubMed PMID: 28326606.

14. Aparicio LF, Jurkovic M, DeLullo J. Decreased bone density in ambulatory patients with duchenne muscular dystrophy. JPediatrOrthop. 2002;22(2):179–81.

15. Crabtree NJ, Roper H, Shaw NJ. Cessation of ambulation results in a dramatic loss of trabecular bone density in boys with Duchenne muscular dystrophy (DMD). Bone. 2022;154:116248. Epub 2021/11/01. doi: 10.1016/j.bone.2021.116248. PubMed PMID: 34718220.

16. Pouwels S, de Boer A, Leufkens HG, Weber WE, Cooper C, van Onzenoort HA, et al. Risk of fracture in patients with muscular dystrophies. Osteoporosis international. 2014;25(2):509–18. Epub 2013/08/21. doi: 10.1007/s00198-013-2442-2. PubMed PMID: 23948807.

17. Laddu DR, Farr JN, Lee VR, Blew RM, Stump C, Houtkooper L, et al. Muscle density predicts changes in bone density and strength: a prospective study in girls. J Musculoskelet Neuronal Interact. 2014;14(2):195–204. Epub 2014/06/01. PubMed PMID: 24879023; PubMed Central PMCID: PMCPMC4414028.

18. Torres-Costoso A, Gracia-Marco L, Sanchez-Lopez M, Garcia-Prieto JC, Garcia-Hermoso A, Diez-Fernandez A, et al. Lean mass as a total mediator of the influence of muscular fitness on bone health in schoolchildren: a mediation analysis. J Sports Sci. 2015;33(8):817–30. Epub 2014/11/12. doi: 10.1080/02640414.2014.964750. PubMed PMID: 25385511.

19. Montgomery E, Pennington C, Isales CM, Hamrick MW. Muscle-bone interactions in dystrophin-deficient and myostatin-deficient mice. Anat Rec A Discov Mol Cell Evol Biol. 2005;286(1):814–22. Epub 2005/08/04. doi: 10.1002/ar.a.20224. PubMed PMID: 16078270.

20. Del Rocio Cruz-Guzman O, Rodriguez-Cruz M, Almeida-Becerril T, Maldonado-Hernandez J, Baeza CW. Muscle function and age are associated with loss of bone mineral density in Duchenne muscular dystrophy. Muscle Nerve. 2019;59(4):417–21. Epub 2019/01/10. doi: 10.1002/mus.26416. PubMed PMID: 30623456.

21. Lott DJ, Taivassalo T, Cooke KD, Park H, Moslemi Z, Batra A, et al. Safety, feasibility, and efficacy of strengthening exercise in Duchenne muscular dystrophy. Muscle Nerve. 2021;63(3):320–6. Epub 2020/12/10. doi: 10.1002/mus.27137. PubMed PMID: 33295018.

22. Rubin C, Turner AS, Bain S, Mallinckrodt C, McLeod K. Anabolism: Low mechanical signals strengthen long bones. Nature. 2001;412(6847):603–4.

23. Rubin CT, Capilla E, Luu YK, Busa B, Crawford H, Nolan DJ, et al. Adipogenesis is inhibited by brief, daily exposure to high-frequency, extremely low-magnitude mechanical signals. ProcNatlAcadSciUSA. 2007;104(45):17879–84.

24. Judex S, Rubin CT. Is bone formation induced by high-frequency mechanical signals modulated by muscle activity? JMusculoskeletNeuronalInteract. 2010;10(1):3–11.

25. Huang RP, Rubin CT, McLeod KJ. Changes in postural muscle dynamics as a function of age. J Gerontol A Biol Sci Med Sci. 1999;54(8):B352–B7.

26. Hannan MT, Cheng DM, Green E, Swift C, Rubin CT, Kiel DP. Establishing the compliance in elderly women for use of a low level mechanical stress device in a clinical osteoporosis study. OsteoporosInt. 2004;15(11):918–26.

27. Gilsanz V, Wren TAL, Sanchez M, Dorey F, Judex S, Rubin C. Low-Level, High-Frequency Mechanical Signals Enhance Musculoskeletal Development of Young Women With Low BMD. Journal of Bone and Mineral Research. 2006;21(9):1464–74. doi: 10.1359/jbmr.060612.

28. Rubin C, Pope M, Chris FJ, Magnusson M, Hansson T, McLeod K. Transmissibility of 15-hertz to 35-hertz vibrations to the human hip and lumbar spine: determining the physiologic feasibility of delivering low-level anabolic mechanical stimuli to skeletal regions at greatest risk of fracture because of osteoporosis. Spine. 2003;28(23):2621–7.

29. Fritton JC, Rubin CT, Qin YX, McLeod KJ. Whole-body vibration in the skeleton: development of a resonance-based testing device. AnnBiomedEng. 1997;25(5):831–9.

30. Chan ME, Uzer G, Rubin CT. The Potential Benefits and Inherent Risks of Vibration as a Non-Drug Therapy for the Prevention and Treatment of Osteoporosis. Curr Osteoporos Rep. 2013. Epub 2013/02/02. doi: 10.1007/s11914-012-0132-1. PubMed PMID: 23371467.

31. Muir J, Kiel DP, Rubin CT. Safety and severity of accelerations delivered from whole body vibration exercise devices to standing adults. Journal of science and medicine in Sports Medicine. 2013. Epub 2013/03/05. doi: 10.1016/j.jsams.2013.01.004. PubMed PMID: 23453990; PubMed Central PMCID: PMC3688642.

32. International Standards O. Evaluation of Human Exposure to Whole-Body Vibration. Geneva: 1985.

33. Bertoli S, Petroni ML, Pagliato E, Mora S, Weber G, Chiumello G, et al. Validation of food frequency questionnaire for assessing dietary macronutrients and calcium intake in Italian children and adolescents. J Pediatr Gastroenterol Nutr. 2005;40(5):555–60. Epub 2005/04/30. doi: 10.1097/01.mpg.0000153004.53610.0e. PubMed PMID: 15861015.

34. Turrini A, Leclercq C, D’Amicis A. Patterns of food and nutrient intakes in Italy and their application to the development of food-based dietary guidelines. Br J Nutr. 1999;81 Suppl 2:S83–9. Epub 2000/09/22. doi: 10.1017/s0007114599001762. PubMed PMID: 10999031.

35. Hangartner TN. Thresholding technique for accurate analysis of density and geometry in QCT, pQCT and microCT images. J Musculoskelet Neuronal Interact. 2007;7(1):9–16. Epub 2007/03/31. PubMed PMID: 17396001.

36. Zemel BS, Leonard MB, Kelly A, Lappe JM, Gilsanz V, Oberfield S, et al. Height adjustment in assessing dual energy x-ray absorptiometry measurements of bone mass and density in children. J Clin Endocrinol Metab. 2010;95(3):1265–73. Epub 2010/01/28. doi: 10.1210/jc.2009-2057. PubMed PMID: 20103654; PubMed Central PMCID: PMCPMC2841534.

37. Taylor A, Konrad PT, Norman ME, Harcke HT. Total body bone mineral density in young children: influence of head bone mineral density. Journal of bone and mineral research. 1997;12(4):652–5. Epub 1997/04/01. doi: 10.1359/jbmr.1997.12.4.652. PubMed PMID: 9101377.

38. Zemel BS, Kalkwarf HJ, Gilsanz V, Lappe JM, Oberfield S, Shepherd JA, et al. Revised reference curves for bone mineral content and areal bone mineral density according to age and sex for black and non-black children: results of the bone mineral density in childhood study. J Clin Endocrinol Metab. 2011;96(10):3160–9. Epub 2011/09/16. doi: 10.1210/jc.2011-1111. PubMed PMID: 21917867; PubMed Central PMCID: PMCPMC3200252.

39. Lee EC, Whitehead AL, Jacques RM, Julious SA. The statistical interpretation of pilot trials: should significance thresholds be reconsidered? BMC Med Res Methodol. 2014;14:41. Epub 2014/03/22. doi: 10.1186/1471-2288-14-41. PubMed PMID: 24650044; PubMed Central PMCID: PMCPMC3994566.

40. Markert CD, Case LE, Carter GT, Furlong PA, Grange RW. Exercise and Duchenne muscular dystrophy: where we have been and where we need to go. Muscle Nerve. 2012;45(5):746–51. Epub 2012/04/14. doi: 10.1002/mus.23244. PubMed PMID: 22499105.

41. Ward K, Alsop C, Caulton J, Rubin C, Adams J, Mughal Z. Low magnitude mechanical loading is osteogenic in children with disabling conditions. JBone MinerRes. 2004;19(3):360–9.

42. Lam TP, Ng BK, Cheung LW, Lee KM, Qin L, Cheng JC. Effect of whole body vibration (WBV) therapy on bone density and bone quality in osteopenic girls with adolescent idiopathic scoliosis: a randomized, controlled trial. Osteoporosis international. 2013;24(5):1623–36. doi: 10.1007/s00198-012-2144-1. PubMed PMID: 23011683.

43. Mogil RJ, Kaste SC, Ferry RJ, Jr., Hudson MM, Mulrooney DA, Howell CR, et al. Effect of Low-Magnitude, High-Frequency Mechanical Stimulation on BMD Among Young Childhood Cancer Survivors: A Randomized Clinical Trial. JAMA oncology. 2016. doi: 10.1001/jamaoncol.2015.6557. PubMed PMID: 26967465.

44. Leonard MB, Shults J, Long J, Baldassano RN, Brown JK, Hommel K, et al. Effect of Low Magnitude Mechanical Stimuli on Bone Density and Structure in Pediatric Crohn’s Disease: A Randomized Placebo Controlled Trial. Journal of bone and mineral research. 2016. doi: 10.1002/jbmr.2799. PubMed PMID: 26821779.

45. Gilsanz V, Wren TA, Sanchez M, Dorey F, Judex S, Rubin C. Low-Level, High-Frequency Mechanical Signals Enhance Musculoskeletal Development of Young Women With Low BMD. JBone MinerRes. 2006;21(9):1464–74.

46. Petryk A, Polgreen LE, Grames M, Lowe DA, Hodges JS, Karachunski P. Feasibility and tolerability of whole-body, low-intensity vibration and its effects on muscle function and bone in patients with dystrophinopathies: a pilot study. Muscle Nerve. 2017;55(6):875–83. Epub 2016/10/09. doi: 10.1002/mus.25431. PubMed PMID: 27718512; PubMed Central PMCID: PMCPMC5385164.

47. Bell JM, Shields MD, Watters J, Hamilton A, Beringer T, Elliott M, et al. Interventions to prevent and treat corticosteroid-induced osteoporosis and prevent osteoporotic fractures in Duchenne muscular dystrophy. Cochrane Database Syst Rev. 2017;1:CD010899. Epub 2017/01/25. doi: 10.1002/14651858.CD010899.pub2. PubMed PMID: 28117876; PubMed Central PMCID: PMCPMC6464928.

48. Joseph S, Wang C, Bushby K, Guglieri M, Horrocks I, Straub V, et al. Fractures and Linear Growth in a Nationwide Cohort of Boys With Duchenne Muscular Dystrophy With and Without Glucocorticoid Treatment: Results From the UK NorthStar Database. JAMA Neurol. 2019;76(6):701–9. Epub 2019/03/12. doi: 10.1001/jamaneurol.2019.0242. PubMed PMID: 30855644; PubMed Central PMCID: PMCPMC6563545.

49. Forbes SC, Arora H, Willcocks RJ, Triplett WT, Rooney WD, Barnard AM, et al. Upper and Lower Extremities in Duchenne Muscular Dystrophy Evaluated with Quantitative MRI and Proton MR Spectroscopy in a Multicenter Cohort. Radiology. 2020;295(3):616–25. Epub 2020/04/15. doi: 10.1148/radiol.2020192210. PubMed PMID: 32286193.

50. Kiel DP, Hannan MT, Barton BA, Bouxsein ML, Sisson E, Lang T, et al. Low-Magnitude Mechanical Stimulation to Improve Bone Density in Persons of Advanced Age: A Randomized, Placebo-Controlled Trial. Journal of bone and mineral research. 2015;30(7):1319–28. doi: 10.1002/jbmr.2448. PubMed PMID: 25581217; PubMed Central PMCID: PMCPMC4834704.

51. Srinivasan S, Agans SC, King KA, Moy NY, Poliachik SL, Gross TS. Enabling bone formation in the aged skeleton via rest-inserted mechanical loading. Bone. 2003;33(6):946–55. Epub 2003/12/18. PubMed PMID: 14678854.

52. Bonewald LF. The amazing osteocyte. Journal of bone and mineral research. 2011;26(2):229–38. Epub 2011/01/22. doi: 10.1002/jbmr.320. PubMed PMID: 21254230.

53. Rubin CT, Bain SD, McLeod KJ. Suppression of the osteogenic response in the aging skeleton. CalcifTissue Int. 1992;50(4):306–13.

54. Rosen CJ, Bouxsein ML. Mechanisms of disease: is osteoporosis the obesity of bone? Nat Clin Pract Rheumatol. 2006;2(1):35–43. doi: 10.1038/ncprheum0070. PubMed PMID: 16932650.

55. Pagnotti GM, Styner M, Uzer G, Patel VS, Wright LE, Ness KK, et al. Combating osteoporosis and obesity with exercise: leveraging cell mechanosensitivity. Nature reviews Endocrinology. 2019;15(6):339–55. Epub 2019/03/01. doi: 10.1038/s41574-019-0170-1. PubMed PMID: 30814687; PubMed Central PMCID: PMCPMC6520125.

56. Soderpalm AC, Kroksmark AK, Magnusson P, Karlsson J, Tulinius M, Swolin-Eide D. Whole body vibration therapy in patients with Duchenne muscular dystrophy--a prospective observational study. J Musculoskelet Neuronal Interact. 2013;13(1):13–8. Epub 2013/03/01. PubMed PMID: 23445910.

57. Vry J, Schubert IJ, Semler O, Haug V, Schonau E, Kirschner J. Whole-body vibration training in children with Duchenne muscular dystrophy and spinal muscular atrophy. Eur J Paediatr Neurol. 2014;18(2):140–9. Epub 2013/10/26. doi: 10.1016/j.ejpn.2013.09.005. PubMed PMID: 24157400.

58. Myers KA, Ramage B, Khan A, Mah JK. Vibration therapy tolerated in children with Duchenne muscular dystrophy: a pilot study. Pediatr Neurol. 2014;51(1):126–9. Epub 2014/05/17. doi: 10.1016/j.pediatrneurol.2014.03.005. PubMed PMID: 24830767.

59. Pel JJ, Bagheri J, van Dam LM, van den Berg-Emons HJ, Horemans HL, Stam HJ, et al. Platform accelerations of three different whole-body vibration devices and the transmission of vertical vibrations to the lower limbs. Med Eng Phys. 2009;31(8):937–44. Epub 2009/06/16. doi: S1350-4533(09)00116-7 [pii] 10.1016/j.medengphy.2009.05.005. PubMed PMID: 19523867.

60. Kiiski J, Heinonen A, Jarvinen TL, Kannus P, Sievanen H. Transmission of vertical whole body vibration to the human body. JBone MinerRes. 2008;23(8):1318–25.

61. Skalsky AJ, Han JJ, Abresch RT, Shin CS, McDonald CM. Assessment of regional body composition with dual-energy X-ray absorptiometry in Duchenne muscular dystrophy: correlation of regional lean mass and quantitative strength. Muscle Nerve. 2009;39(5):647–51. Epub 2009/04/07. doi: 10.1002/mus.21212. PubMed PMID: 19347922.

62. Forbes SC, Willcocks RJ, Triplett WT, Rooney WD, Lott DJ, Wang DJ, et al. Magnetic resonance imaging and spectroscopy assessment of lower extremity skeletal muscles in boys with Duchenne muscular dystrophy: a multicenter cross sectional study. PLoS One. 2014;9(9):e106435. Epub 2014/09/10. doi: 10.1371/journal.pone.0106435. PubMed PMID: 25203313; PubMed Central PMCID: PMCPMC4159278.

63. Willcocks RJ, Forbes SC, Walter GA, Sweeney L, Rodino-Klapac LR, Mendell JR, et al. Assessment of rAAVrh.74.MHCK7.micro-dystrophin Gene Therapy Using Magnetic Resonance Imaging in Children With Duchenne Muscular Dystrophy. JAMA Netw Open. 2021;4(1):e2031851. Epub 2021/01/05. doi: 10.1001/jamanetworkopen.2020.31851. PubMed PMID: 33394000; PubMed Central PMCID: PMCPMC7783546.

64. Manson JE, Skerrett PJ, Greenland P, VanItallie TB. The escalating pandemics of obesity and sedentary lifestyle. A call to action for clinicians. Arch Intern Med. 2004;164(3):249–58. doi: 10.1001/archinte.164.3.249. PubMed PMID: 14769621.

65. Schara U, Mortier Mortier W. Long-Term Steroid Therapy in Duchenne Muscular Dystrophy-Positive Results versus Side Effects. J Clin Neuromuscul Dis. 2001;2(4):179–83. Epub 2001/06/01. doi: 10.1097/00131402-200106000-00002. PubMed PMID: 19078632.

66. Seibel MJ, Cooper MS, Zhou H. Glucocorticoid-induced osteoporosis: mechanisms, management, and future perspectives. Lancet Diabetes Endocrinol. 2013;1(1):59–70. Epub 2014/03/14. doi: 10.1016/S2213-8587(13)70045-7. PubMed PMID: 24622268.

67. Luu YK, Capilla E, Rosen CJ, Gilsanz V, Pessin JE, Judex S, et al. Mechanical stimulation of mesenchymal stem cell proliferation and differentiation promotes osteogenesis while preventing dietary-induced obesity. Journal of bone and mineral research. 2009;24(1):50–61. Epub 2008/08/22. doi: 10.1359/jbmr.080817. PubMed PMID: 18715135; PubMed Central PMCID: PMCPmc2689082.

68. Al Wren T, Lee DC, Kay RM, Dorey FJ, Gilsanz V. Bone density and size in ambulatory children with cerebral palsy. Dev Med Child Neurol. 2011;53(2):137–41. Epub 2010/12/21. doi: 10.1111/j.1469-8749.2010.03852.x. PubMed PMID: 21166671; PubMed Central PMCID: PMCPMC3064513.

69. Rajapakse CS, Johncola AJ, Batzdorf AS, Jones BC, Al Mukaddam M, Sexton K, et al. Effect of Low-Intensity Vibration on Bone Strength, Microstructure, and Adiposity in Pre-Osteoporotic Postmenopausal Women: A Randomized Placebo-Controlled Trial. Journal of bone and mineral research. 2021;36(4):673–84. Epub 2020/12/15. doi: 10.1002/jbmr.4229. PubMed PMID: 33314313.

70. Chang G, Rajapakse CS, Regatte RR, Babb J, Saxena A, Belmont HM, et al. 3 Tesla MRI detects deterioration in proximal femur microarchitecture and strength in long-term glucocorticoid users compared with controls. Journal of magnetic resonance imaging. 2015;42(6):1489–96. Epub 2015/06/16. doi: 10.1002/jmri.24927. PubMed PMID: 26073878; PubMed Central PMCID: PMCPMC4676948.

71. Arpan I, Willcocks RJ, Forbes SC, Finkel RS, Lott DJ, Rooney WD, et al. Examination of effects of corticosteroids on skeletal muscles of boys with DMD using MRI and MRS. Neurology. 2014;83(11):974–80. Epub 2014/08/08. doi: 10.1212/WNL.0000000000000775. PubMed PMID: 25098537; PubMed Central PMCID: PMCPMC4162304.

72. Ozcivici E, Luu YK, Adler B, Qin YX, Rubin J, Judex S, et al. Mechanical signals as anabolic agents in bone. NatRevRheumatol. 2010;6(1):50–9.

73. Xie L, Jacobson JM, Choi ES, Busa B, Donahue LR, Miller LM, et al. Low-level mechanical vibrations can influence bone resorption and bone formation in the growing skeleton. Bone. 2006;39(5):1059–66. Epub 2006/07/11. doi: 10.1016/j.bone.2006.05.012. PubMed PMID: 16824816.

74. Mettlach G, Polo-Parada L, Peca L, Rubin CT, Plattner F, Bibb JA. Enhancement of neuromuscular dynamics and strength behavior using extremely low magnitude mechanical signals in mice. J Biomech. 2014;47(1):162–7. Epub 2013/10/26. doi: 10.1016/j.jbiomech.2013.09.024. PubMed PMID: 24157062; PubMed Central PMCID: PMCPMC3881264.

75. Frechette DM, Krishnamoorthy D, Adler BJ, Chan ME, Rubin CT. Diminished satellite cells and elevated adipogenic gene expression in muscle as caused by ovariectomy are averted by low-magnitude mechanical signals. Journal of applied physiology. 2015;119(1):27–36. Epub 2015/05/02. doi: 10.1152/japplphysiol.01020.2014. PubMed PMID: 25930028; PubMed Central PMCID: PMCPmc4491530.

76. Batra A, Harrington A, Lott DJ, Willcocks R, Senesac CR, McGehee W, et al. Two-Year Longitudinal Changes in Lower Limb Strength and Its Relation to Loss in Function in a Large Cohort of Patients With Duchenne Muscular Dystrophy. Am J Phys Med Rehabil. 2018;97(10):734–40. Epub 2018/05/08. doi: 10.1097/PHM.0000000000000957. PubMed PMID: 29734234; PubMed Central PMCID: PMCPMC6148402.

